# Overview of the 8^th^ Social Media Mining for Health Applications (#SMM4H) Shared Tasks at the AMIA 2023 Annual Symposium

**DOI:** 10.1101/2023.11.06.23298168

**Authors:** Ari Z. Klein, Juan M. Banda, Yuting Guo, Ana Lucia Schmidt, Dongfang Xu, Jesus Ivan Flores Amaro, Raul Rodriguez-Esteban, Abeed Sarker, Graciela Gonzalez-Hernandez

## Abstract

The aim of the Social Media Mining for Health Applications (#SMM4H) shared tasks is to take a community-driven approach to address the natural language processing and machine learning challenges inherent to utilizing social media data for health informatics. The eighth iteration of the #SMM4H shared tasks was hosted at the AMIA 2023 Annual Symposium and consisted of five tasks that represented various social media platforms (Twitter and Reddit), languages (English and Spanish), methods (binary classification, multi-class classification, extraction, and normalization), and topics (COVID-19, therapies, social anxiety disorder, and adverse drug events). In total, 29 teams registered, representing 18 countries. In this paper, we present the annotated corpora, a technical summary of the systems, and the performance results. In general, the top-performing systems used deep neural network architectures based on pre-trained transformer models. In particular, the top-performing systems for the classification tasks were based on single models that were pre-trained on social media corpora. To facilitate future work, the datasets—a total of 61,353 posts—will remain available by request, and the CodaLab sites will remain active for a post-evaluation phase.

## INTRODUCTION

With more than 70% of adults in the United States[1] and nearly 60% of people worldwide[2] using social media, the aim of the Social Media Mining for Health Applications (#SMM4H) shared tasks is to take a community-driven approach to address the natural language processing (NLP) and machine learning challenges inherent to utilizing the vast amount of data on social media for health informatics. The eighth iteration of the #SMM4H shared tasks was hosted at the American Medical Informatics Association (AMIA) 2023 Annual Symposium and consisted of five tasks. Task 1 was a binary classification task to distinguish English-language tweets that self-report a COVID-19 diagnosis from those that do not.[3] Task 2 was a multi-class classification task to categorize users’ sentiment toward therapies in English-language tweets as *positive, negative*, or *neutral*.[4] Task 3 was a sequence labeling task to extract COVID-19 symptoms in tweets written in Latin American Spanish. Task 4 was a binary classification task to distinguish English-language Reddit posts that self-report a social anxiety disorder diagnosis from those that do not. Task 5 was a normalization task to map adverse drug events (ADEs) in English-language tweets to their standard concept ID in the MedDRA vocabulary.[5]

Teams could register for a single task or multiple tasks. In total, 29 teams registered, representing 18 countries. Teams were provided with gold standard annotated training and validation sets to develop their systems and, subsequently, a blind test set for the final evaluation. After receiving the test set, teams were given five days to submit the predictions of their systems to CodaLab[6-10]—a platform that facilitates data science competitions—for automatic evaluation, promoting the systematic comparison of performance. Among the 29 teams that registered, 17 teams submitted at least one set of predictions: 8 teams for Task 1, 6 teams for Task 2, 2 teams for Task 3, 8 teams for Task 4, and 3 teams for Task 5. Teams were then invited to submit a short manuscript describing their system, and 12 of the 17 teams did. Each of these 12 system descriptions was peer-reviewed by at least two reviewers. In this paper, we present the annotated corpora, a technical summary of the systems, and the performance results, providing insights into state-of-the-art methods for mining social media data for health informatics.

## METHODS

### Data collection

We collected a total of 61,353 social media posts for the five tasks. For Task 1, the dataset included 10,000 English-language tweets that mentioned a personal reference to the user and keywords related to both COVID-19 and a positive test, diagnosis, or hospitalization.[3] For Task 2, the dataset included 5364 English-language tweets that mentioned a total of 32 therapies, such as medication, behavioral, and physical.[4] These tweets were posted by a cohort of users who self-reported chronic pain on Twitter,[11] so it is likely that the sentiments associated with the therapies are being expressed by patients who are actually experiencing them. For Task 3, the dataset included 10,150 tweets that were written in Latin American Spanish and reported COVID-19 symptoms of the user or someone known to the user, building on a previous iteration of this task involving the multi-class classification of Spanish-language tweets that mentioned COVID-19 symptoms.[12] For Task 4, the dataset included 5140 English-language Reddit posts in the *r/socialanxiety* subreddit, written by users aged 13-25 years. For Task 5, the dataset included 29,449 English-language tweets that mentioned medications, with many from previous iterations of this task.[12-17]

### Annotation

For all five tasks, at least a subset of the social media posts was annotated by multiple annotators. Table 1 presents inter-annotator agreement (IAA) and the distribution of the posts in the training, validation, and test sets. For Task 1, 1728 (17.3%) of the tweets were labeled as self-reporting a COVID-19 diagnosis—a positive test, clinical diagnosis, or hospitalization—and 8272 (82.7%) as not.[3] For Task 2, 998 (18.6%) of the tweets were labeled as *positive* sentiment toward a therpay, 619 (11.5%) as *negative* sentiment, and 3747 (69.9%) as *neutral* sentiment.[4] For Task 3, the spans of text containing COVID-19 symptoms were annotated by medical doctors who are native speakers of Latin American Spanish. Tweets were included more than once if they mentioned multiple symptoms, with a single symptom annotated for each instance. For Task 4, 2428 (38%) of the Reddit posts were labeled as self-reporting a confirmed or probable clinical social anxiety disorder diagnosis and 3962 (62%) as not. For Task 5, 3021 (10.3%) of the tweets were annotated with a span of text containing an ADE and a corresponding MedDRA ID, and 26,428 (89.7%) were not. Among the 1224 tweets in the test set that reported an ADE, 272 (22.2%) of them reported an ADE that was not in the training or validation sets.

**Table 1.** Inter-annotator agreement (IAA) and distribution of social media posts in the training, validation, and test sets for the five #SMM4H 2023 shared tasks.

| Task | Platform | Language | Training | Validation | Test | Total | IAA |
| --- | --- | --- | --- | --- | --- | --- | --- |
| 1 | Twitter | English | 7600 | 400 | 2000 | 10,000 | 0.79<sup>a</sup> |
| 2 | Twitter | English | 3009 | 753 | 1602 | 5364 | 0.82<sup>b</sup> |
| 3 | Twitter | Spanish | 6021 | 1979 | 2150 | 10,150 | 0.88<sup>b</sup> |
| 4 | Reddit | English | 4500 | 640 | 1250 | 6390 | 0.80<sup>b</sup> |
| 5 | Twitter | English | 17,385 | 915 | 11,149 | 29,449 | 0.68<sup>c</sup> |

<sup>a</sup> Fleiss' kappa
<sup>b</sup> Cohen's kappa
<sup>c</sup> F<sub>1</sub>-score

### Classification

For Task 1, the baseline[3] and five of the seven teams (Table 2 in Results) that submitted a system description (Shayona, UQ,[18] IICU-DSRG, KUL, and TMN[19]) used a classifier based on COVID-Twitter-BERT—a transformer model pre-trained on tweets related to COVID-19.[20] Two of these five teams used additional BERT-based models for the feature representation (IICU-DSRG) or ensemble learning (TMN[19]); IICU-DSRG used BioMed-RoBERTa-Base,[21] and TMN[19] used RoBERTa-Large[22] and Twitter-RoBERTa-Base.[23] One of the two teams that did not use COVID-Twitter-BERT[20] (Explorers) used an ensemble of RoBERTa-Base,[22] CPM-RoBERTa, and BERTweet[24] models, following 5-fold cross-validation for each individual model. They also used the tweets in the Task 1 and Task 4 training and validation sets for continued domain-adaptive pretraining of these models.[21] The other team (BFCI) used a Passive Aggressive classifier with bigrams as features in a bag-of-words representation. Three of the teams (UQ,[18] KUL, and Explorers) used techniques for addressing the class imbalance, including data augmentation (UQ[18] and KUL), over-/under-sampling (UQ[18]), and class weights (UQ[18] and Explorers).

**Table 2.** Performance of 18 teams’ systems on the test sets for the five #SMM4H 2023 shared tasks.

| Team | Task 1<sup>a</sup> | Task 2<sup>b</sup> | Task 3<sup>c</sup> | Task 4<sup>d</sup> | Task 5a<sup>e</sup> | Task 5b<sup>f</sup> |
| --- | --- | --- | --- | --- | --- | --- |
| ABCD | - | - | - | - | 0.188 | 0.089 |
| BFCI | 0.637 | 0.714 | - | - | - | - |
| CEN | - | - | - | 0.718 | - | - |
| DS4DH | - | - | - | - | **0.426** | **0.292** |
| Explorers | 0.872 | - | **0.94** | 0.695 | - | - |
| HULAT-UC3M | - | 0.669 | - | - | - | - |
| I2R-MI | - | 0.752 | - | - | 0.322 | 0.195 |
| IICU-DSRG | 0.898 | - | - | - | - | - |
| ITT | - | - | - | 0.728 | - | - |
| KUL | 0.877 | - | - | - | - | - |
| Mantis | - | - | - | 0.838 | - | - |
| MUCS | 0.741 | 0.185 | - | - | - | - |
| Shayona | **0.943** | - | - | 0.810 | - | - |
| ThaparUni | - | - | - | 0.842 | - | - |
| TMN | 0.845 | - | - | - | - | - |
| UQ | 0.943 | **0.778** | - | **0.871** | - | - |
| Unknown-1<sup>g</sup> | - | 0.695 | - | 0.841 | - | - |
| Unknown-2<sup>g</sup> | - | - | 0.89 | - | - | - |

<sup>a</sup> F<sub>1</sub>-score for the class of tweets that self-reported a COVID-19 diagnosis
<sup>b</sup> Micro-averaged F<sub>1</sub>-score for all three classes of tweets
<sup>c</sup> Strict F<sub>1</sub>-score for identifying the character offsets of COVID-19 symptoms
<sup>d</sup> F<sub>1</sub>-score for the class of Reddit posts that self-reported a social anxiety disorder diagnosis
<sup>e</sup> Micro-averaged F<sub>1</sub>-score for all tweets with MedDRA IDs
<sup>f</sup> Micro-averaged F<sub>1</sub>-score for tweets with MedDRA IDs that were not seen during training
<sup>g</sup> CodaLab submission could not be cross-referenced with a registered team name

For Task 2, two of the four teams that submitted a system description (UQ[18] and I2R-MI) used BERT-based classifiers: BERTweet-Large[24] (UQ[18]) and RoBERTa-Base[22] (IR2-MI). These teams also used techniques for addressing the class imbalance, including data augmentation (UQ[18]), over-sampling (UQ[18]), under-sampling (UQ[18] and IR2-MI), and class weights (UQ[18]). Furthermore, UQ[18] used a large language model (LLM), GPT-3.5,[25] to verify the predictions of the BERTweet-Large[24] classifier, automatically changing the prediction to the majority (i.e., *neutral* sentiment) class if the LLM did not find evidence in the tweet to support the classification of *positive* or *negative* sentiment. The two teams that did not use a BERT-based classifier (BFCI and HULAT-UC3M) used a Multilayer Perceptron classifier with 5-fold cross-validation (BFCI) and a Random Forest classifier with feature engineering (HULAT-UC3M).

For Task 4, all five of the teams that submitted a system description (UQ,[18] ThaparUni, Mantis, Shayona, and Explorers) used BERT-based models, with two of the teams (UQ[18] and Explorers) using BERTweet[24] models. As in Task 2, UQ[18] used techniques for addressing the class imbalance and a LLM to correct the predictions of the BERTweet[24] classifier. As in Task 1, Explorers used BERTweet[24] in an ensemble with RoBERTa-Base[22] and CPM-RoBERTa models, domain-adaptive pretraining,[21] and techniques for addressing the class imbalance. Also as in Task 1, Shayona used a COVID-Twitter-BERT[20] model with gradient boosting.[26] Along with Explorers, the other two teams (ThaparUni and Mantis) used an ensemble of pre-trained transformer models; while ThaparUni used RoBERTa,[22] ERNIE 2.0,[27] and XLNet[28] models, which were pre-trained on general-domain corpora, Mantis used MentalRoBERTa[29] and PyschBERT[30] models, which were pre-trained on corpora related to mental health, including Reddit posts.

### Extraction

For Task 3, the one team that submitted a system description (Explorers) used the W^2^NER framework[31] for named entity recognition and an ensemble of three Spanish BERT-based models: Spanish BERT (BETO),[32] BETO+NER, and a version of BETO that was fine-tuned using the Spanish portion of the XLNI corpus.[33] As in Task 1 and Task 4, they also used the tweets in the training and validation sets for continued domain-adaptive pretraining[21] of these models. Two baselines used BETO[32] and COVID-Twitter-BERT[20] models. For Task 5, the baseline[4] and two teams that submitted a system description (DS4DH[34] and I2R-MI) also used pre-trained transformer models, in a pipeline approach that involves extracting ADEs as input for mapping the tweets to MedDRA terms: BERT[35] (baseline[5]), BERTweet[24] (DS4DH[34]), and T5[36] (I2R-MI).

### Normalization

For Task 5, the two teams that submitted a system description (DS4DH[34] and I2R-MI) took similar approaches to ADE normalization, using similarity metrics to compare the vector representations of extracted ADEs to MedDRA terms, based on pre-trained sentence transformer models. DS4DH[34] used multiple sentence transformers to represent each extracted ADE—S-PubMedBERT,[37] All-MPNet-Base-v2,[38] All-DistilRoBERTa-v1,[38] All-MiniLM-L6-v2,[38] and a custom sentence transformer that integrates knowledge from the Unified Medical Language System (UMLS) Metathesaurus—and aggregated their similarity scores via reciprocal-rank fusion,[39] while I2R-MI used only a single SBERT[38] model for the similarity ranking.

In contrast, the baseline[5] took a learning-based approach, using a BERT[35] model for multi-class classification.

## RESULTS AND DISCUSSION

Table 2 presents the performance of 18 teams’ systems on the test sets for the five tasks. For Task 1, the evaluation metric was the F_1_-score for the class of tweets that self-reported a COVID-19 diagnosis. Shayona and UQ[18] achieved nearly identical F_1_-scores (0.943) using a COVID-Twitter-BERT[20] model, with Shayona marginally outperforming UQ;[18] however, neither gradient boosting[26] (Shayona) nor techniques for addressing the class imbalance (UQ[18]) substantially improved performance over a COVID-Twitter-BERT baseline classifier[3] (0.938), which was trained using the *Flair* library with 5 epochs, a batch size of 8, and a learning rate of 1e-5. While the recall of Shayona’s (0.938) and UQ’s[18] (0.950) systems outperformed that of the baseline (0.914), the baseline achieved the highest precision (0.962) among all of the systems. These three systems outperformed IICU-DSRG’s and TMN’s[19] systems that used COVID-Twitter-BERT[20] with additional BERT-based models. While KUL did not use additional models, the lower performance of their system (0.877) may be a result of the variation in hyperparameters used for fine-tuning COVID-Twitter-BERT[20]: 10 epochs, a batch size of 32, and a learning rate of 5e-5. All of the systems that used BERT-based classifiers substantially outperformed BFCI’s Passive Aggressive classifier (0.637).

For Task 2, the evaluation metric was the micro-averaged F_1_-score for all three classes of tweets: *positive* sentiment, *negative* sentiment, and *neutral* sentiment. UQ[18] achieved not only the highest micro-averaged F_1_-score (0.778), but also the highest F_1_-score for each of the *positive* (0.611), *negative* (0.415), and *neutral* (0.859) classes. An ablation study by UQ[18] showed that the use of a LLM[25] to verify the predictions of the BERTweet-Large[24] classifier improved the micro-averaged F_1_-score by more than three points, contributing to outperforming I2R-MI’s RoBERTa-Base[22] classifier (0.752). UQ’s[18] and I2R-MI’s BERT-based classifiers outperformed BFCI’s Multilayer Perceptron (0.714) and HULAT-UC3M’s Random Forest (0.669) classifiers.

For Task 3, the evaluation metric was the strict F_1_-score for identifying the character offsets of COVID-19 symptoms. Explorers’ ensemble of three Spanish BERT-based models and continued domain-adaptive pretraining[21] of these models achieved not only a higher F_1_-score (0.94) than that of BETO[31] (0.90) and COVID-Twitter-BERT[20] (0.92) baselines, but also a higher precision (0.94) and recall (0.93). Despite the high performance of Explorers’ system, their error analysis revealed that colloquial/regional spelling variants of symptoms remain a challenge; for example, their system was able to identify *gripe*, but not *gripa* and *gripes*.

For Task 4, the evaluation metric was the F_1_-score for the class of Reddit posts that self-reported a social anxiety disorder diagnosis. Using the same methods as in Task 2, UQ[18] also achieved the highest F_1_-score (0.871) for Task 4. As with UQ,[18] Explorers also used BERTweet[24] and techniques for addressing the class imbalance, though they used it an ensemble with two additional BERT-based models and achieved a substantially lower F_1_-score (0.695). ThaparUni’s ensemble of three transformer models that were pre-trained on general-domain corpora (0.842)— RoBERTa,[22] ERNIE 2.0,[27] and XLNet[28]—marginally outperformed Mantis’ ensemble of two transformer models that were pre-trained on corpora related to mental health, including Reddit posts (0.838)—MentalRoBERTa[29] and PyschBERT.[30] Both of these ensemble-based systems outperformed Shayona’s COVID-Twitter-BERT[20] classifier (0.810), suggesting that this model may not generalize to non-COVID-19 diagnoses.

To enable novel approaches for Task 5, in contrast to previous iterations of this task that used a pipeline-based evaluation,[12-17] the evaluation did not include the output of classification or extraction. The primary evaluation metric was the micro-averaged F_1_-score for all tweets in the test set with MedDRA IDs—in other words, for all tweets that reported an ADE. A secondary evaluation metric was based on a zero-shot learning setup—that is, for MedDRA IDs in the test set that were not seen during training. While DS4DH[34] achieved the highest overall precision (0.449) and I2R-MI achieved the highest recall for the unseen ADEs (0.406), the baseline[5] achieved the highest overall recall (0.508) and F_1_-score (0.452) and the highest precision (0.335) and F_1_-score (0.363) for the unseen ADEs. In contrast to DS4DH’s[34] and I2R-MI’s two-component pipelines, the baseline’s pipeline includes an initial binary classification component to detect tweets that mention an ADE, pre-filtering the tweets for downstream extraction and normalization.[5]

In general, the top-performing team for each of the five tasks used a deep neural network architecture based on pre-trained transformer models. In total, 10 of the 12 teams that submitted a system description used pre-trained transformer models. In particular, the top-performing team for each of the three classification tasks used a model that was pre-trained on a social media corpus. While half—five of the ten—teams that participated in the classification tasks used an ensemble-based system, the top-performing system for each of these three tasks was based on a single model. In addition, the techniques that four of these ten teams used for addressing the class imbalance did not appear to substantially improve performance. In contrast, for the second iteration of the #SMM4H shared tasks, which was hosted at the AMIA 2017 Annual Symposium, deep neural networks were used in only approximately half of the systems and were outperformed by Support Vector Machine (SVM) and Logistic Regression classifiers on highly imbalanced data.[17]

## CONCLUSION

This paper presented an overview of the #SMM4H 2023 shared tasks, which represented various social media platforms (Twitter and Reddit), languages (English and Spanish), methods (binary classification, multi-class classification, extraction, and normalization), and topics (COVID-19, therapies, social anxiety disorder, and adverse drug events). To facilitate future work, the datasets will remain available by request, and the CodaLab sites[6-10] will remain active to automatically evaluate new systems against the blind test sets, promoting the ongoing systematic comparison of performance.

## Data Availability

According to the Twitter Terms of Service, the content (e.g., text) of Tweet Objects cannot be made publicly available; however, a limited number of Tweet Objects are permitted to be shared directly. Requests for data can be sent to Ari Z. Klein or Graciela Gonzalez-Hernandez.

## AUTHOR CONTRIBUTIONS

AZK: data collection, annotation, baseline system, system description review, and wrote the manuscript. JMB: shared task conceptualization, data collection, annotation, baseline system, CodaLab site, system description review, and wrote the manuscript. YG: data collection, annotation, CodaLab site, system description review, and wrote the manuscript. ALS: data collection, annotation, CodaLab site, system description review, and wrote the manuscript. DX: data collection, annotation, baseline system, CodaLab site, system description review, and wrote the manuscript. JIFA: data collection, CodaLab site, and edited the manuscript. RRE: shared task conceptualization and edited the manuscript. AS: shared task conceptualization and edited the manuscript. GGH: shared task conceptualization and guidance, and edited the manuscript.

## ACKNOWLEDGMENTS

The authors thank those who contributed to annotating the data, the program committee of the #SMM4H 2023 Workshop, and additional peer reviewers of the system description papers.

